# Denoising Longitudinal Social Media for Pandemic Monitoring

**DOI:** 10.1101/2024.06.29.24309690

**Authors:** Shixu Lin, Lucas Garay, Yining Hua, Zhijiang Guo, Xiaolin Xu, Jie Yang

## Abstract

**Objective:** Current studies leveraging social media data for disease monitoring face challenges like noisy colloquial language and insufficient tracking of user disease progression in longitudinal data settings. This study aims to develop a pipeline for collecting, cleaning, and analyzing large-scale longitudinal social media data for disease monitoring, with a focus on COVID-19 pandemic.

**Materials and Methods:** This pipeline initiates by screening COVID-19 cases from tweets spanning February 1, 2020, to April 30, 2022. Longitudinal data is collected for each patient, two months before and three months after self-reporting. Symptoms are extracted using Name Entity Recognition (NER), followed by denoising with a combination of Graph Convolutional Network (GCN) and Bidirectional Encoder Representations from Transformers (BERT) model to retain only User Symptom Mentions (USM). Subsequently, symptoms are mapped to standardized medical concepts using the Unified Medical Language System (UMLS). Finally, this study conducts symptom pattern analysis and visualization to illustrate temporal changes in symptom prevalence and co-occurrence.

**Results:** This study identified 191,096 self-reported COVID-19-positive cases from COVID-19-related tweets and retrospectively collected 811,398,280 historical tweets, of which 2,120,964 contained symptoms information. After denoising, 39% (832,287) of symptom-sharing tweets reflected user-related mentions. The trained USM model achieved an F1 score of 0.926. Further analysis revealed a higher prevalence of upper respiratory tract symptoms during the Omicron period compared to the Delta and wild-type periods. Additionally, there was a pronounced co-occurrence of lower respiratory tract and nervous system symptoms in the wild-type strain and Delta variant.

**Conclusion:** This study established a robust framework for pandemic monitoring via social media, integrating denoising of user-related symptom mentions and longitudinal data. The findings underscore the importance of denoising procedures in revealing accurate prevalence trends, thereby minimizing biases in symptom analysis.

## Introduction

Social media platforms bring together a large number of users for health-related discussions, such as disease symptoms sharing, preventive measures discussion, and health information dissemination, which have shown great potential in the development and evaluation of infectious disease outbreaks, health crisis management, and health promotion strategies [1–3]. During the COVID-19 pandemic, social media has been utilized in public health for various purposes such as monitoring disease outbreaks [4], establishing early warning systems [5–8], gathering public opinions regarding medications and vaccines [9–12], and evaluating mental health [13]. Furthermore, social media has the potential to monitor disease symptoms in populations [14–16], and identify specific cohorts for observational studies [17–19]. Data from social media is low-cost, wide-coverage, and real-time. This provides unprecedented opportunities for syndromic surveillance when compared to traditional bio-surveillance which relies on phone surveys, electronic health records, and laboratory testing, which are time-consuming, restricted access and limited population coverage[20–22].

The challenges in social media analytics, particularly in the context of public health concepts, stem from the limitations of traditional natural language processing (NLP) techniques that heavily rely on rule-based approaches [23]. These traditional methods have shown poor performance in the social media domain due to the noisy colloquial language used, leading to a shift towards machine-learning-based approaches in recent years [24]. The informal and unstructured nature of social media language complicates the accurate capture of essential public health concepts. This challenge is exacerbated by the fact that many symptom terms identified through these methods may not necessarily be user-related content but could be news reports or general discussions about health events [25–27]. The accuracy of symptom identification and classification is paramount for ensuring the integrity of individual analysis, as misclassification of posts and inaccurate reporting on user-related health events can lower the confidence of subsequent data analysis [28].

While many studies have focused on cross-sectional analyses, there is a growing recognition of the need for more in-depth and continuous tracking of users over time [10,29–31]. Longitudinal social media analysis is essential for comprehending the dynamics of user’s health status during the pandemic, particularly concerning public health issues such as disease evolution and user behaviors [32,33]. Understanding these dynamics over time is essential for developing targeted interventions to control diseases, such as vaccine advertisement, public health education of infectious disease [34]. By tracking these trends longitudinally, researchers can identify patterns and associations that may inform public health measures aimed at promoting healthier behaviors during crises like the COVID-19 pandemic.

The COVID-19 pandemic is one of the major pandemics throughout documented human history, with various variants emerging and affecting different regions [35]. These variants, such as the Omicron variant, have led to changes in the frequency and severity of common symptoms like fever, cough, sore throat, and fatigue [36]. Understanding the impact of these variants is crucial for public health responses and policymaking. One critical aspect of COVID-19 is the persistence of symptoms in some individuals, leading to what is commonly referred to as long COVID. Research has shown that persistent symptoms after a mild COVID-19 infection can have major consequences for work and daily functioning [37]. Furthermore, the burden of post-acute COVID-19 symptoms is substantial, with a high percentage of individuals reporting persistent symptoms [38]. These persistent symptoms can be associated with the severity of the initial COVID-19 infection [39]. The broad impact of COVID-19, the emergence of variants, the persistence of symptoms in long COVID, and the significance of analyzing symptom persistence are crucial areas of research that require further investigation to enhance our understanding of the disease and improve patient outcomes.

While current studies leveraging social media data for disease symptom tracking have made meaningful progress, they often face challenges such as insufficient long-term individual tracking or limited noise reduction, which reduces the accuracy of the results [10,14–16,29–31]. In this work, we proposed a pipeline that introduces a comprehensive NLP based framework that denoises longitudinal social media for pandemic monitoring. The key components of this pipeline include patient screening and retrospective data collection, symptom identification, symptom denoising, and symptom normalization. Taken COVID-19 as the use case, we demonstrated how this pipeline can support downstream processes related to tracking and analyzing the evolution of symptom patterns during different pandemic phases. The three main contributions of this study are:

1. By denoising the dataset, we provided clean and reliable symptom data for analysis, ensuring that downstream tasks are based on accurate information.
2. We integrated longitudinal data in the analysis which offers a dynamic perspective on how symptoms manifest and evolve throughout different stages of the pandemic.
3. We established a unified framework that enhances the process and interpretation of data for pandemic monitoring.

## Methods

### Overall Workflow

The overall workflow is visualized in Figure 1 and consists of two main parts. The first part comprises four key modules: patient screening, symptom identification, symptom denoising, and symptom mapping. These modules work in tandem to filter, identify, refine, and categorize symptom data from social media, ensuring the extraction of high-quality, relevant health information. The second part focuses on evaluating symptom prevalence, exploring risk recovery times, and assessing the strength of symptom co-occurrence across different variants. By integrating these components, our pipeline offers a robust tool for long-term monitoring and analysis of symptom patterns, illustrated using COVID-19 as a case study.

**Figure 1.**
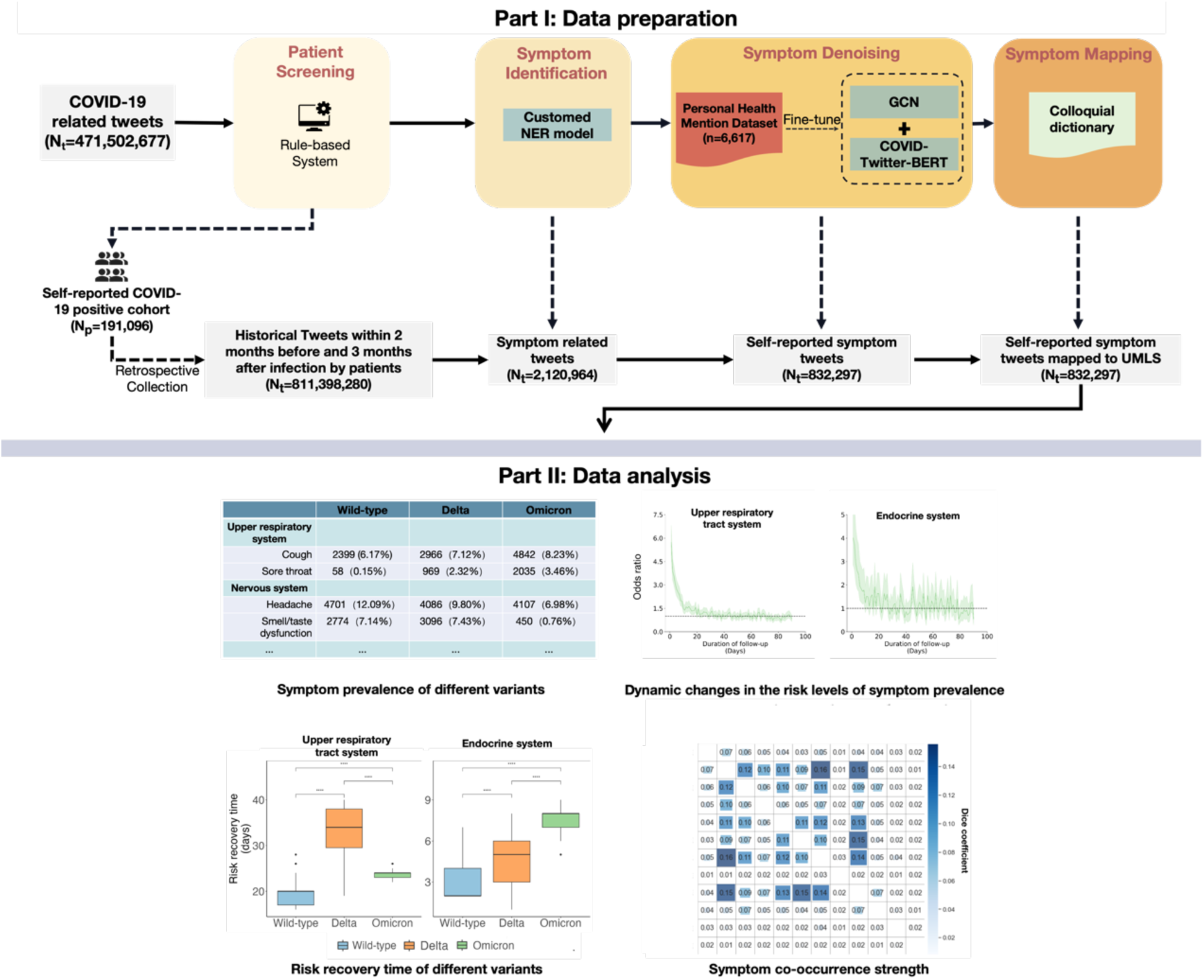
Overall workflow. (Note: N_t_, N_p_, NER, and GCN represent the number of tweets, the number of patients, Name Entity Recognition, and Graph Convolutional Network, respectively)

### Data Collection

Using Twitter’s Application Programming Interface (API), we downloaded non-retweeted English tweets related to COVID-19 from an open-source database of COVID-19 tweets [40,41], accessed through a collaborative network at Harvard Medical School. The collection period for these tweets spanned from February 1, 2020, to April 30, 2022. These tweets were selected based on their inclusion of popular topics or specific keywords associated with COVID-19, such as ‘COVID-19’ and ‘SARS-CoV-2’.

### Patient screening

Using predefined rules, we filtered self-reported COVID-19-positive cases (Figure 2). These rules include constructing a list of keywords and phrases, applying dependency parsing techniques, and filtering through regular expressions. Initially, we created a list of keywords and phrases directly related to self-reported COVID-19-positive cases, focusing on identifying tweets with expressions like “get COVID-19” and “test positive.” Subsequently, we employed dependency parsing [42] to determine the structural relationships among sentence elements, such as subjects and objects, to identify subjects associated with specific COVID-19-related expressions. For example, in tweets like “I got COVID” and “I tested positive,” dependency parsing confirmed that ‘I,’ as the subject, was directly associated with these expressions. Finally, we used a unique form of regular expression, known as zero-width assertions [43], to identify texts near specific words or phrases (such as personal pronouns), excluding tweets that, despite containing keywords, were contextually incorrect, such as “I wonder if I got COVID” and “I imagine I tested positive.” In these cases, the personal pronoun ‘I’ does not actually indicate a positive COVID-19 result.

**Figure 2.**
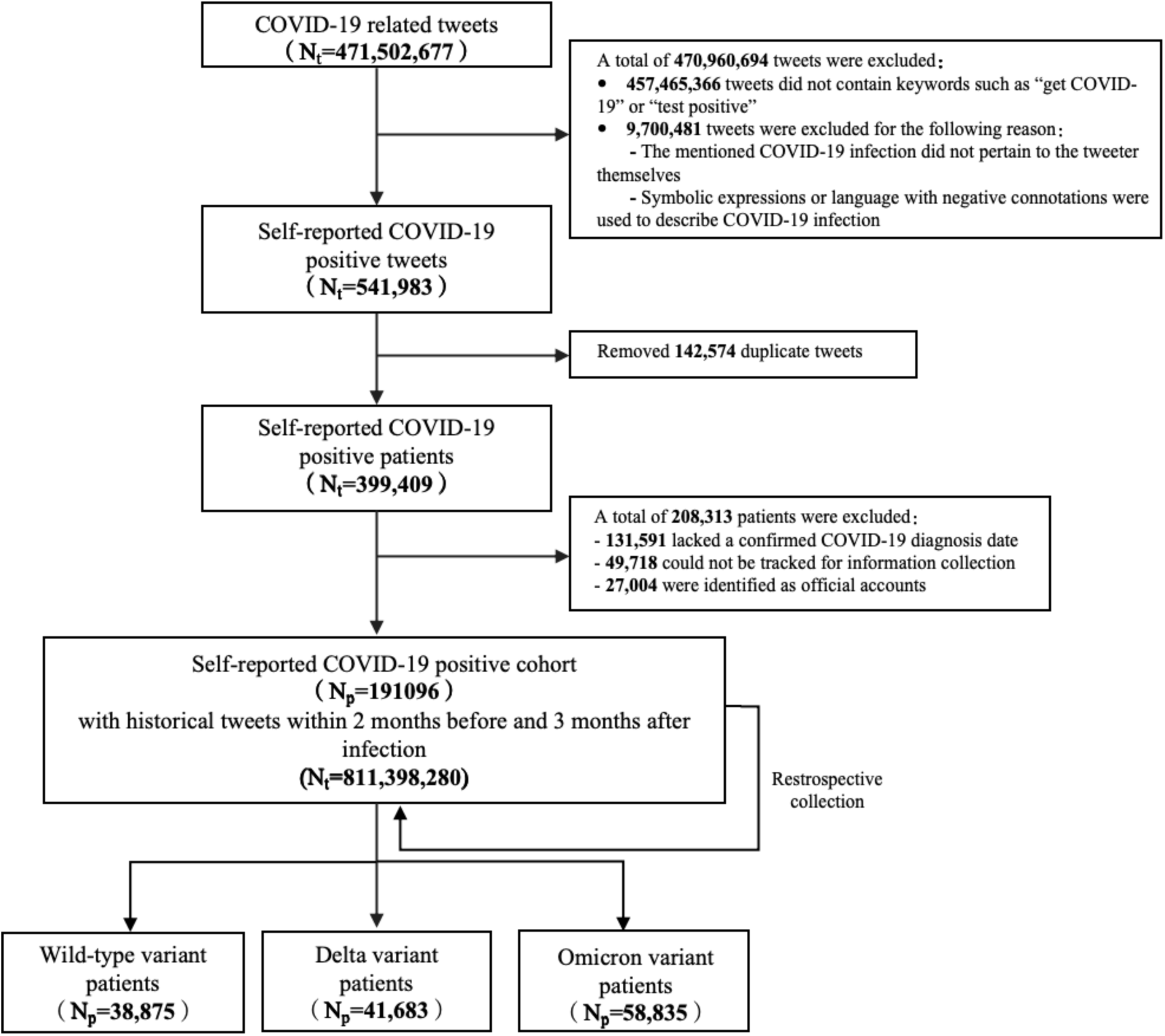
Flow diagram for the step-by-step COVID-19 patient screening. (Note: N_t_ represents the number of tweets, and Np represents the number of patients.)

Upon completion of these steps, tweets from individuals self-reporting as COVID-19 positive were collated. Subsequently, we removed duplicate tweets by using unique user IDs, retaining only the first tweet in which a user self-reports being positive for COVID-19. At this point, each self-reporting COVID-19-positive user was associated with their first self-reporting tweet, which included a timestamp indicating when the tweet was posted. However, it is important to note that this timestamp does not directly reflect the time of self-reported COVID-19-positive. Therefore, to infer the time of self-reported COVID-19-positive, we used regular expressions to extract time-related information (such as “yesterday,” “last Sunday,” etc.) from the positive tweets and combined this information with the tweet’s timestamp. For example, if a user posted “I got COVID-19 yesterday” on “2020-03-05,” the inferred date of self-reported positivity would be “2020-03-04.” This study retained only the data of users whose self-reported COVID-19-positive dates could be precisely determined. Additionally, the standard method M3 [44] was employed to identify users’ organizational identities and filter out official accounts, ensuring the research primarily focuses on content generated by individual users. After successfully filtering positive patients, we conducted a retrospective data collection for each patient, including historical tweet data from two months before to three months after the time of self-reported COVID-19-positive.

In the subsequent analysis of symptom patterns, we categorized patients into three mutually exclusive groups based on the time of self-reported COVID-19-positive: the Wild-type group, the Delta group, and the Omicron group. The Wild-type group includes patients who reported positive from April 27, 2020 to December 21, 2020; the Delta group includes those reporting positive from June 5, 2021 to November 22, 2021; and the Omicron group covers patients reporting positive from December 20, 2021 to April 30, 2022. These time frames were chosen based on the periods during which each target variant strain accounted for more than 80% of all sequences [45].

### Symptom Identification

To extract symptom information from historical self-reported COVID-19 user data, we employed a Named Entity Recognition (NER) model to identify COVID-19 symptom entities. The NER model utilized in the study was CT-BERT (COVID-Twitter-BERT) [46]. This model underwent training on the METS-CoV (Medical Entities and Targeted Sentiments on COVID-19-related Tweets) dataset within the YATO framework [47,48].

### Symptom Denoising

Given that symptom entities identified by the NER model may not accurately reflect users’ actual health conditions, we developed the User Symptom Mention (USM) text classification model. This model aims to determine whether symptoms mentioned in social media tweets genuinely represent the health issues experienced by the users.

To train the USM classification model, we developed a USM dataset (Table 1), collaboratively annotated by team members with medical backgrounds. The dataset facilitates a comprehensive examination through binary classification of whether the symptoms discussed are directly pertinent to the users themselves. It comprises 3,000 tweets encompassing 6,617 symptom entities, with the annotation process divided into three stages (The statistics and distribution of USM dataset are listed in Table 2). The F1 score was used as the primary metric to assess the consistency among annotators. All annotators worked on the same corpus and strictly adhered to the annotation guidelines. After completing the annotations, the project supervisor compared all results to establish the final gold standards, which were then used to calculate annotator consistency. This process resulted in F1 scores of 0.805, 0.835, and 0.864 for the three rounds of annotations, respectively, underscoring the reliability of our method. The USM dataset’s train-dev-test splitting is with a ratio of 70:15:15.

**Table 1.** USM label categories and examples.

| Category | Example |
| --- | --- |
| User-related-symptom | “I woke up yesterday at 3 am in a **cold sweat**.”<br>“I got a really bad cramp in my foot.”<br>“My back just started hurting.” |
| Non-user-related-symptom | “It’s SO cold!!! My poor chihuahua just **shivers**.”<br>“It’s interesting to read your point, but I’m **feeling sick to my stomach**.”<br>“A Month Before a **Heart Attack**, Your Body Will Warn You With These 8 Signals.” |

**Table 2.** Summary statistics of the USM dataset.

| Symptom category | Number of user-related symptom<br>entities | Number of non-user-related<br>symptom entities | Total |
| --- | --- | --- | --- |
| Circulatory system | 133 | 93 | 226 |
| Digestive system | 316 | 353 | 669 |
| Endocrine system | 54 | 28 | 82 |
| General and others | 494 | 537 | 1031 |
| Genitourinary system | 34 | 54 | 88 |
| Immune system | 29 | 79 | 108 |
| Integumentary system | 174 | 124 | 298 |
| Lower respiratory tract system | 359 | 398 | 757 |
| Musculoskeletal system | 265 | 251 | 516 |
| Nervous system | 1038 | 1030 | 2068 |
| Psychiatric system | 241 | 187 | 428 |
| Upper respiratory tract system | 200 | 146 | 346 |
| Total | 3314 | 3303 | 6617 |

The text classification model [49] integrates the complex structural and relational processing capabilities of Graph Convolutional Networks (GCN) [50] with the deep semantic understanding of the CT-BERT model. Initially, the model employs the CT-BERT text encoder to extract textual features, yielding an embedded representation of the text. Concurrently, it utilizes the “stanza” library [51] to extract a sentence dependency graph from the original text, capturing syntactic information and dependencies between words to enhance the understanding of semantic nuances and contextual meanings. GCNs associate syntactically related words with the target aspect and, by learning through GCN layers, improve the model’s comprehension of textual structures and semantics by incorporating distant word relationships and syntactic information. The embedded text representation and the dependency graph are input together into the GCN layer, merging deep semantic and syntactic structural information to generate new features. These features, along with the dependency graph, are then fed into subsequent GCN layers for further feature fusion.

After processing through the GCN layers, the extracted features are combined with the initial text embedding. This combination is then input into an attention mechanism layer, which assigns attention scores based on semantic features closely related to the target vocabulary—specifically, symptom-related words—within the hidden state vectors. Finally, a fully connected layer utilizes these attention-weighted features to output the ultimate classification results (Figure 3).

**Figure 3.**
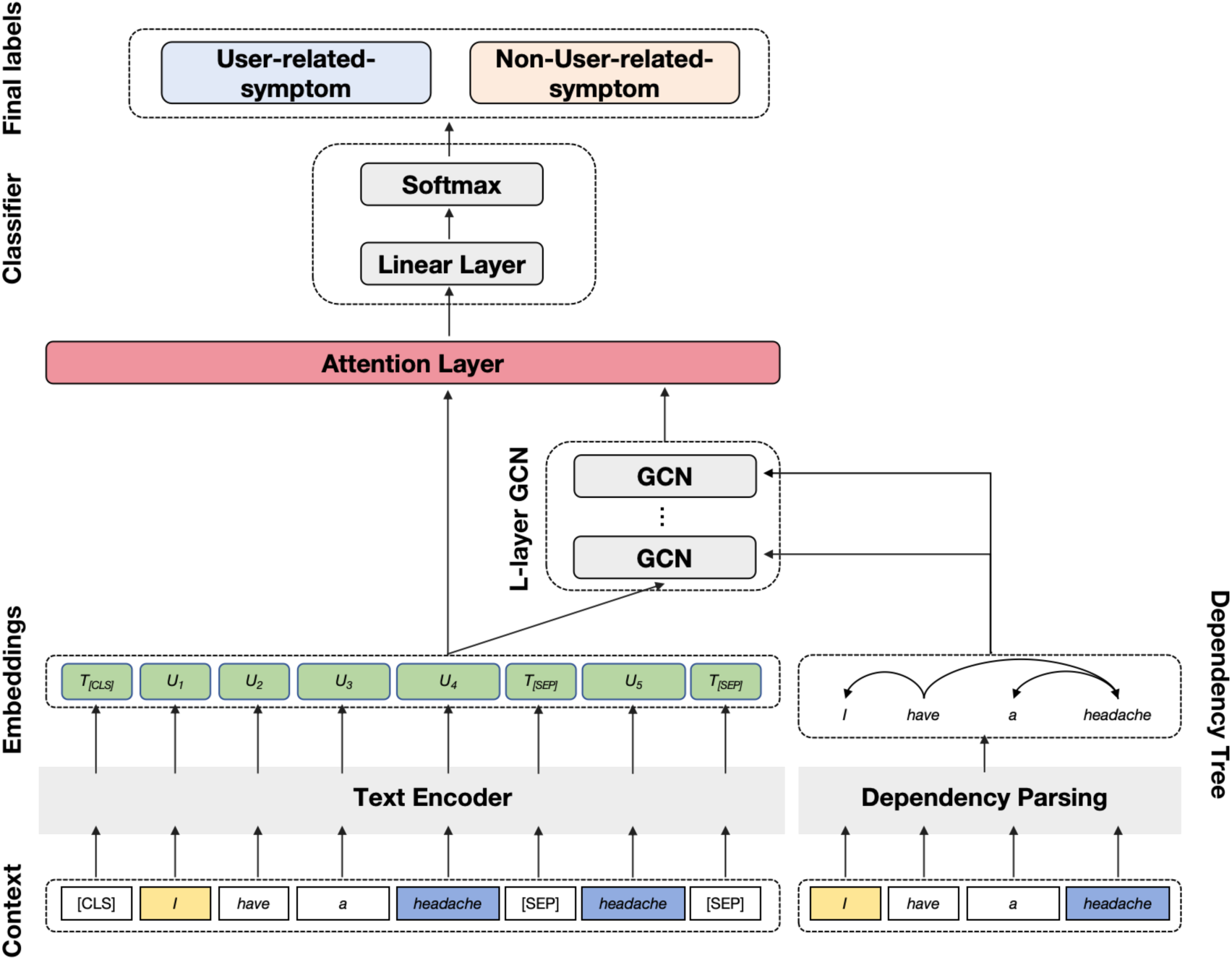
Overall structure of the USM classification model. (Note: [CLS], [SEP], and GCN represent classifier token, sentence separator, and Graph Convolutional Networks, respectively)

### Symptom Mapping

Given the informal nature of symptom descriptions in social media text data, it is necessary to standardize the mapping of colloquial expressions to unified symptom concepts for statistical analysis. Manual construction of a comprehensive lexicon is impractical. Therefore, our previous work [31] integrates a normalization and mapping module that utilizes the Unified Medical Language System (UMLS) [52] to create a colloquial dictionary. We further categorized these concepts into twelve primary categories based on physiological systems, including a psychiatric system for mental symptoms. For greater precision, respiratory symptoms were subdivided into upper and lower respiratory tract systems. Concepts that did not fit into these categories were placed in a ‘general and others’ category.

### Symptom Pattern

The subsequent statistical analyses were conducted using Python version 3.6.3, along with the Lifelines and Statsmodels packages. All Latent Class Analysis (LCA) models were implemented using Mplus version 8.3.

#### Symptom Prevalence

We analyzed the frequency and prevalence rates of each symptom category within the 90 days following the date of self-reported COVID-19 positive outcome. This examination was conducted across different periods characterized by dominant variants of the virus. By focusing on these variant-specific periods, our analysis provides insights into how symptom prevalence varies with different viral strains, further demonstrating the robustness of our pipeline in tracking and categorizing symptoms accurately over time.

#### Dynamic Changes in Symptom Prevalence Risk

The daily Odds Ratio (OR) was employed to track changes in symptom prevalence post-SARS-CoV-2 infection. This ratio is calculated by dividing the daily symptom prevalence by the baseline prevalence. The baseline period, defined as 60 to 30 days before the date of self-reported COVID-19-positive, was selected as a reference point to reflect the normal prevalence level of symptoms in the patient population prior to COVID-19 infection. OR value greater than 1 indicates an increased risk of symptom prevalence; OR value of 1 indicates a risk level at the normal prevalence level. To address the variability of daily OR values, we performed 100 non-parametric bootstrap resamplings of daily symptom prevalence rates to estimate confidence intervals for daily OR values. The risk resolution period was defined as the point when the OR and its confidence interval first dropped to 1. The range of the risk resolution period was also determined through 100 bootstrap iterations. To describe the range of risk resolution period, we used the median and interquartile range (Q1-Q3). The Wilcoxon rank-sum test was used to compare medians.

#### Symptom Co-occurrence

We analyzed the co-occurrence strength of symptoms across 12 symptom categories. Co-occurrence refers to the simultaneous presence of two symptom categories in the same patient within 0 to 90 days after the self-reported COVID-19 positive date. To calculate the strength of symptom co-occurrence, we employed the Kaplan-Meier method to estimate the probability of symptom A occurring between two time points (t_1_ to t_2_) [53].

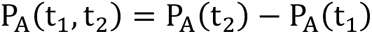

P_A_(t) represents the probability of symptom A occurring up to time t since the start of the follow-up period. In estimating probabilities using the Kaplan-Meier method, only the first occurrence of an event within the follow-up period is considered. Thus, if a patient exhibits the same symptom multiple times, only the first occurrence is included in the analysis.

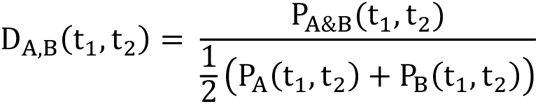

Similarly, the co-occurrence probability of symptoms A and B, P_A&B_ (t_1_, t_2_), is estimated using the Kaplan-Meier method. Using these probability estimates, we calculate the Dice coefficient [54] to measure the degree of simultaneous occurrence of two symptoms. The Dice coefficient ranges from 0 to 1, with values closer to 1 indicating a higher degree of symptom co-occurrence.

## Results

As shown in Figure 2, a total of 191,096 self-reported COVID-19-positive patients were screened from COVID-19-related tweets. We retrospectively collected 811,398,280 historical tweets from these patients. Using the NER model, we identified and filtered 2,120,964 tweets containing symptom information. Further denoising through the USM model filtered out 832,297 tweets documenting the patients’ own symptom information, excluding 1,288,667 (60.76%) non-USM tweets. The remaining data were then utilized for the analysis of symptom patterns.

### The USM Dataset and Model Performance

We developed the USM dataset (Table 2) for model training and evaluation. The dataset encompasses 6,617 symptom entities with a broad range of coverage. Neurological symptoms are the most prevalent, totaling 2,068 entities, while endocrine system symptoms are the least common, with only 82 entities. Overall, the ratio of user-related symptoms to non-user-related symptoms in the dataset is nearly 1:1. However, within the immune system category, the proportion of non-user-related symptom entities is notably higher, reaching 73.15% (79 out of 108).

Table 3 compares the performance of our developed model (GCN+BERT) with several baseline models, including Logistic Regression (LR), Random Forest (RF), Support Vector Machine (SVM), and another BERT-based model (BERT-SPC). Our GCN+BERT model outperforms the baseline models in all three metrics: F1 score, precision, and recall. Specifically, the GCN+BERT model achieves an F1 score of 0.926, precision of 0.910, and recall of 0.950. In contrast, the LR and RF models both have an F1 score of 0.850, with precision and recall scores slightly above 0.850. The SVM model performs similarly with an F1 score of 0.845. The BERT-SPC model, while demonstrating improved performance over the traditional classifiers with an F1 score of 0.912, still falls short compared to our GCN+BERT model. These results highlight the performance of our GCN+BERT model in accurately classifying user-related symptom mentions, effectively leveraging both graph convolutional networks and BERT for enhanced contextual understanding and classification accuracy. As a result, we selected GCN+BERT as our USM model in text classification tasks.

**Table 3.** Performance Comparison of Different Classifiers on USM Dataset.

| Classifier | F1 | Precision | Recall |
| --- | --- | --- | --- |
| LR | 0.850 | 0.845 | 0.853 |
| RF | 0.850 | 0.852 | 0.854 |
| SVM | 0.845 | 0.842 | 0.845 |
| BERT-SPC | 0.912 | 0.910 | 0.925 |
| GCN+BERT | 0.926 | 0.910 | 0.950 |

Table 4 shows the performance of the USM model in classifying health status mentions (including user-related symptoms and non-user-related symptoms) across different physiological systems. Overall, the USM model demonstrates high performance, with an average F1 score of 0.926, average precision of 0.921, and average recall of 0.930. These results indicate the USM model’s high accuracy and reliability in differentiating between user-related symptoms and non-user-related symptoms, effectively filtering and categorizing health-related information from social media texts.

**Table 4.**
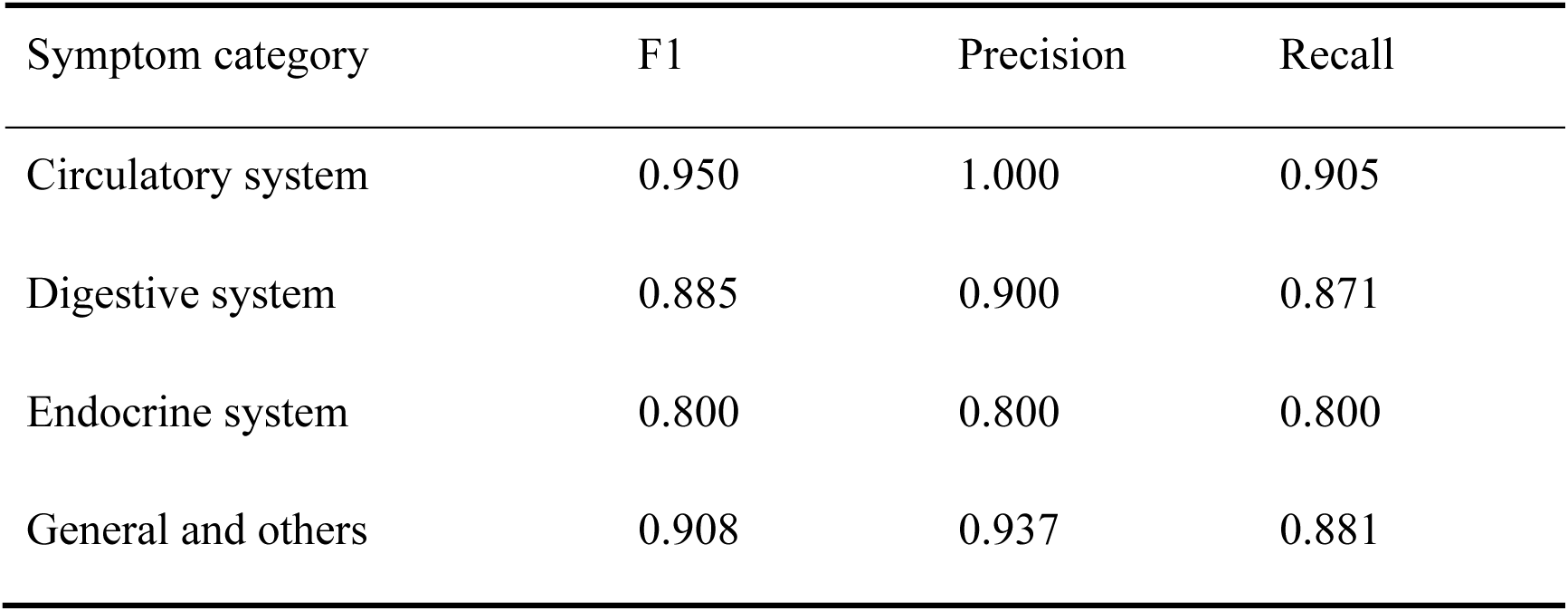
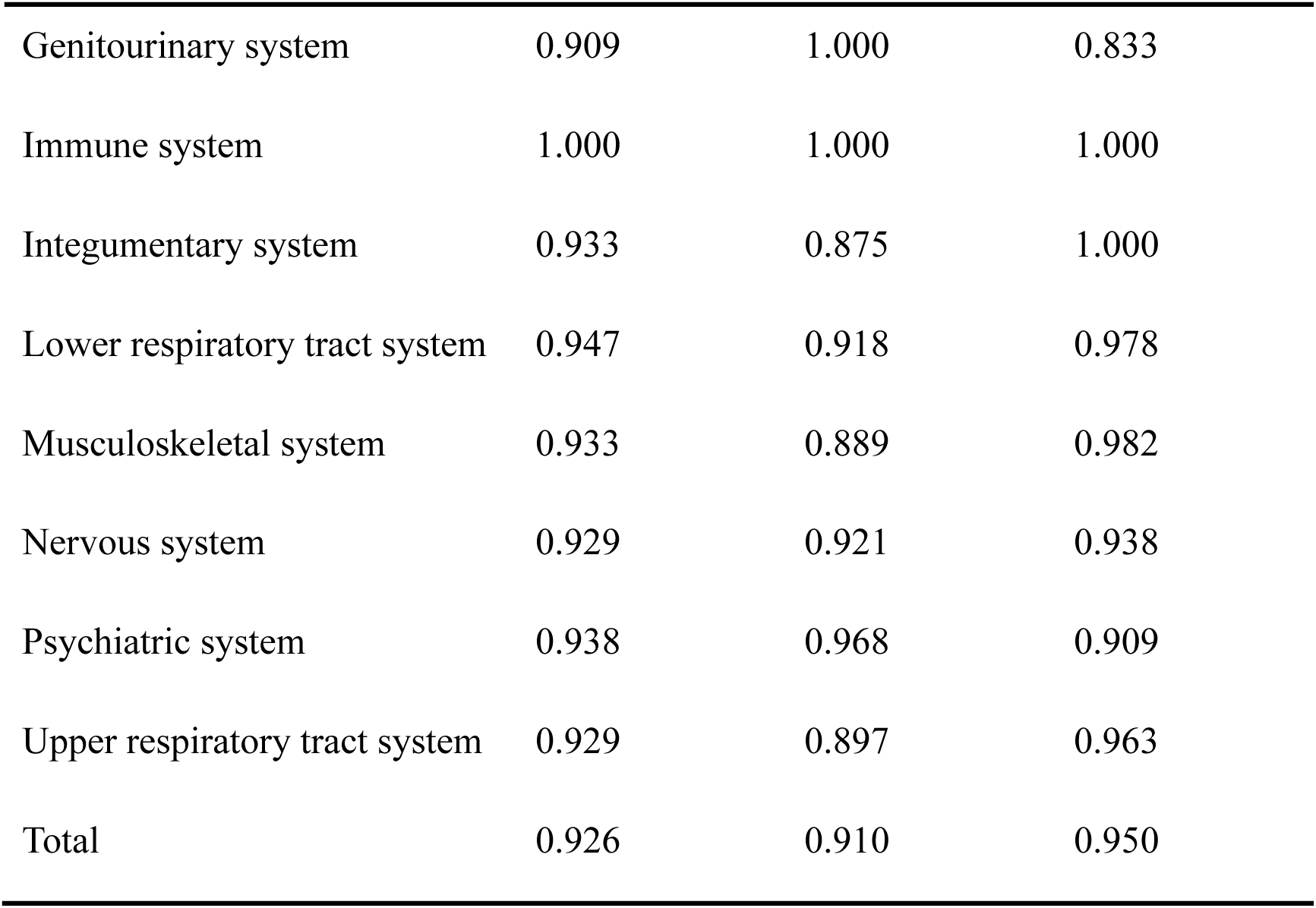
Performance of the USM model on the test set.

### Symptom prevalence among different variants

We observed that, as the variants evolved, there was a substantial decrease in the occurrence rates of most symptoms, with this decline being most pronounced in the Omicron variant. This trend was particularly noticeable in symptoms related to the nervous system, psychiatric system, musculoskeletal system, lower respiratory system, and immune system (Table 5).

**Table 5.** Frequency and prevalence of common symptoms in patients with different variants.

|  | Wild-type<br>(n=38875) | Delta<br>(n=41683) | Omicron<br>(n=58835) | Total<br>Population<br>(n=191096) |
| --- | --- | --- | --- | --- |
| **Circulatory system** |  |  |  |  |
| Swelling | 1178(3.03) | 989(2.37) | 674(1.15) | 4388(2.30) |
| Palpitations | 736(1.89) | 493(1.18) | 364(0.62) | 2412(1.26) |
| Chest pain | 736(1.89) | 453(1.09) | 339(0.58) | 2274(1.19) |
| Chest tightness | 687(1.77) | 393(0.46) | 145(0.25) | 1883(0.99) |
| Easy bruising | 632(1.63) | 439(1.05) | 49(0.08) | 1844(0.96) |
| **Digestive system** |  |  |  |  |
| Nausea and/or vomiting | 1546(3.98) | 1214(2.91) | 1501(2.55) | 6124(3.20) |
| Abdominal pain | 1304(3.35) | 964(2.31) | 337(0.57) | 3992(2.09) |
| Loss of appetite | 560(1.44) | 444(1.07) | 90(0.15) | 1722(0.90) |
| **Endocrine system** |  |  |  |  |
| Sweating | 542(1.39) | 471(1.13) | 487(0.83) | 2197(1.15) |
| Hair loss | 385(0.99) | 251(0.60) | 72(0.12) | 1039(0.54) |
| **General and others** |  |  |  |  |
| Fever | 2273(5.85) | 2286(5.48) | 2158(3.67) | 9924(5.19) |
| Chills | 1290(3.32) | 1219(2.92) | 1103(1.87) | 5620(2.94) |
| Cold sweat | 678(1.74) | 1055(2.53) | 1832(3.11) | 4724(2.47) |
| **Genitourinary system** |  |  |  |  |
| Hematuria | 519(1.34) | 516(1.24) | 434(0.74) | 2139(1.12) |
| Dysmenorrhea | 35(0.09) | 39(0.09) | 97(0.16) | 226(0.12) |
| **Immune system** |  |  |  |  |
| Anaphylaxis | 1300(3.34) | 1019(2.44) | 514(0.87) | 4635(2.43) |
| **Integumentary system** |  |  |  |  |
| Skin lesion | 2249(5.79) | 2371(5.69) | 2117(3.60) | 10421(5.45) |
| Rash | 1099(2.83) | 770(1.85) | 658(1.12) | 3951(2.07) |
| Flushing | 635(1.63) | 485(1.16) | 512(0.87) | 2432(1.27) |
| **Lower respiratory tract system** |  |  |  |  |
| Shortness of breath | 1913(4.92) | 1185(2.84) | 772(1.31) | 5726(3.00) |
| Wheezing | 886(2.28) | 586(1.41) | 177(0.30) | 2513(1.32) |
| Cyanosis | 461(1.19) | 351(0.84) | 297(0.50) | 1660(0.87) |
| **Musculoskeletal system** |  |  |  |  |
| Pain | 4488(11.54) | 3614(8.67) | 1762(2.99) | 15420(8.07) |
| Back pain | 1785(4.69) | 1271(3.05) | 292(0.50) | 5459(2.86) |
| Muscle cramps | 1494(3.84) | 1148(2.75) | 548(0.93) | 4984(2.61) |
| Nervous system |  |  |  |  |
| Headaches | 4701(12.09) | 4086(9.80) | 4107(6.98) | 18961(9.92) |
| Fatigue | 4445(11.43) | 4085(9.80) | 4020(6.83) | 18601(9.73) |
| Problem with smell or taste | 2774(7.14) | 3096(7.43) | 450(0.76) | 9699(5.08) |
| Insomnia | 2382(6.13) | 1582(3.80) | 485(0.82) | 7038(3.68) |
| Psychiatric system |  |  |  |  |
| Anxiety | 3030(7.79) | 1752(4.20) | 377(0.64) | 8240(4.31) |
| Psychosis | 2969(7.64) | 3171(7.61) | 2837(4.82) | 13014(6.81) |
| Depression | 1980(5.09) | 1371(3.29) | 862(1.47) | 6404(3.35) |
| Upper respiratory tract system |  |  |  |  |
| Cough | 2399(6.17) | 2966(7.12) | 4842(8.23) | 13534(7.08) |
| Pain in throat | 58(0.15) | 969(2.32) | 2035(3.46) | 3930(2.06) |
| Nosebleed | 474(1.22) | 959(2.30) | 1842(3.13) | 4441(2.32) |
| Itchy throat | 295(0.76) | 1087(2.61) | 1782(3.03) | 4188(2.19) |
| Sinonasal congestion | 575(1.48) | 768(1.84) | 1000(1.70) | 3117(1.63) |

In symptoms related to the nervous system, the prevalence of anosmia or ageusia, headache, fatigue, and insomnia largely decreased from 7.14%, 12.09%, 11.43%, and 6.13% in the wild-type variant to 0.76%, 6.98%, 6.83%, and 0.82% in the Omicron variant, respectively. Additionally, the proportions of psychiatric symptoms such as anxiety and depression decreased from 7.79% and 5.09% in the wild-type variant to 0.64% and 1.47% in the Omicron variant, respectively. This indicates that as the virus strain evolves, there may be an improvement in the neurological and mental health conditions of patients.

In the musculoskeletal system, the prevalence of muscle pain, back pain, and muscle cramps decreased from 11.54%, 4.69%, and 3.84% in the wild-type variant to 2.99%, 0.50%, and 0.93% in the Omicron variant, respectively, reflecting a substantial reduction in musculoskeletal symptoms. Similarly, the prevalence of dyspnea in the lower respiratory system decreased from 4.92% in the wild-type variant to 1.31% in the Omicron variant. The prevalence of immune responses in the immune system substantially decreased from 3.34% to 0.87%. In the digestive system, the proportion of abdominal pain symptoms decreased from 3.35% to 0.57%. In the circulatory system, the proportions of edema, chest tightness, and chest pain symptoms decreased from 3.03%, 1.89%, and 1.77% to 1.15%, 0.25%, and 0.58%, respectively. These reductions indicate that the impact of the Omicron variant strain on multiple body systems has been reduced.

Notably, certain symptoms such as cold sweats increased in prevalence from 1.74% in the wild-type variant to 3.11% in the Omicron variant. Additionally, symptoms of the upper respiratory system such as cough, sore throat, nasal bleeding, itchy throat, and nasal congestion also showed varying degrees of increase, rising from 6.17%, 0.15%, 1.22%, 0.76%, and 1.48% to 8.23%, 3.46%, 3.13%, 3.03%, and 1.70%, respectively. This may reflect a more pronounced impact of the Omicron variant strain on the upper respiratory tract.

### Changes in symptom prevalence risk among different variants

Figure 4 depicts the changing pattern of symptom prevalence risk over time in the COVID-19-positive population. As time progresses from the point of self-reported infection, the daily Odds Ratio (OR) shows a gradual declining trend, eventually stabilizing near the baseline level. This trend suggests that, over time, the risk of symptom prevalence in the population will gradually return to normal levels. However, there are differences in the rate of decline and the time to return to normal levels (hereinafter referred to as ‘risk resolution period’) for different physiological systems. The risk resolution period for the immune system is shown to be 6 days (Q1-Q3, 5–7 days), for the endocrine system 8 days (Q1-Q3, 8–10 days), while the risk resolution period for the lower respiratory system is 32 days (Q1-Q3, 32–33 days), and for the nervous system, it extends up to 38 days (Q1-Q3, 35–38 days).

**Figure 4.**
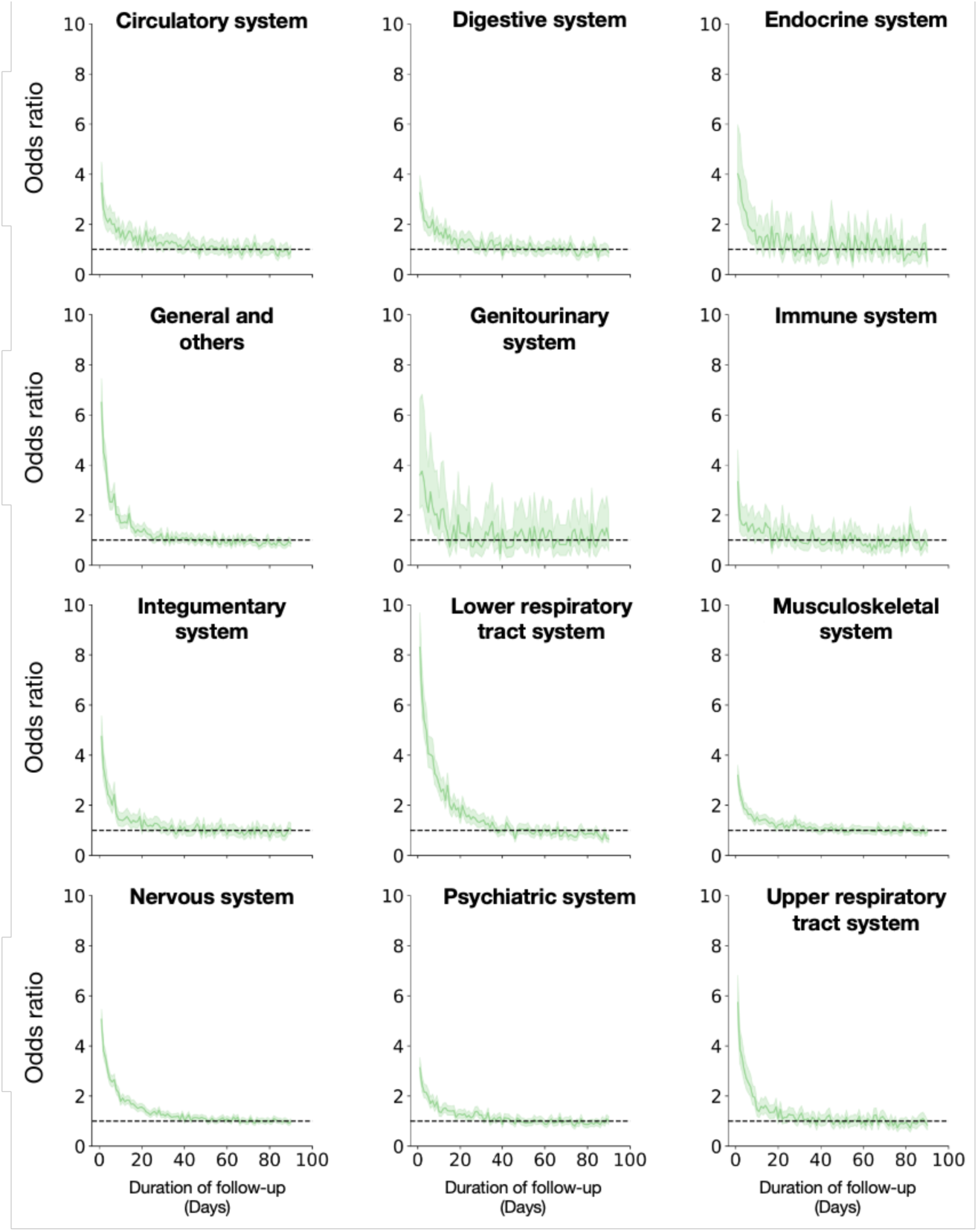
Changes in the prevalence risk of different physiological systems among the self-report COVID-19-positive cohort. (Note: The shaded area represents the 95% confidence interval, and the dashed line indicates the normal level of symptom prevalence risk.)

Among different variants, the risk resolution period for the same physiological system varies (Figure 5). Specifically, in the lower respiratory system, the risk resolution period for the Delta variant group is 34 days (Q1-Q3, 30–38 days), significantly longer than the 20 days (Q1-Q3, 17–20 days) for the wild-type strain group and 24 days (Q1-Q3, 23–24 days) for the Omicron variant group (*P*<0.001). In the integumentary system, the risk resolution period for the Delta variant group is 11 days (Q1-Q3, 11–14 days), compared to 8 days (Q1-Q3, 6–8 days) for the wild-type strain group and 8 days (Q1-Q3, 8–8 days) for the Omicron variant group (*P*<0.001). In the endocrine system, the risk resolution period for the Omicron variant group is 8 days (Q1-Q3, 7–8 days), whereas it is 2 days (Q1-Q3, 2–4 days) for the wild-type strain group and 5 days (Q1-Q3, 3–6 days) for the Delta variant group (*P*<0.001). For other physiological systems, the differences in risk resolution period are less pronounced.

**Figure 5.**
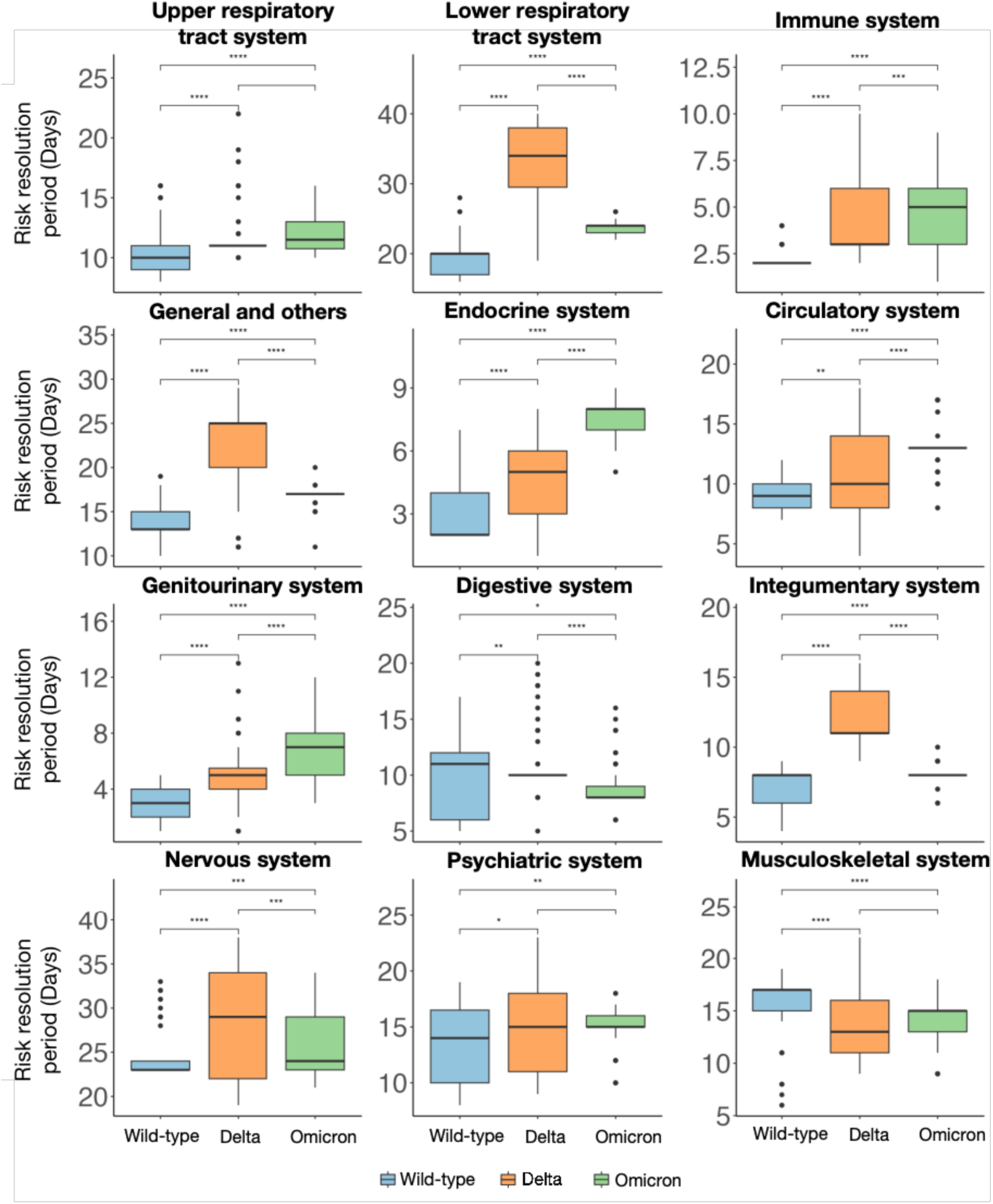
Comparative distribution of risk resolution period across physiological systems with different variants. (Note: Significance levels are marked as: \**P*<0.05, \*\**P*<0.01, \*\*\**P*<0.001, \*\*\*\**P*<0.0001.)

### Symptom Co-occurrence of SARS-CoV-2 Strains among different variants

In the wild-type strain, there is a relatively strong co-occurrence between the nervous system and the lower respiratory system with other systems. The strongest co-occurrence is between the nervous system and general and others, with a Dice coefficient of 0.16. Additionally, the co-occurrence between the lower respiratory system and the nervous system, as well as between the lower respiratory system and the upper respiratory system, also show a higher degree of closeness, with Dice coefficients of 0.15 each. In the Delta variant, although the overall co-occurrence pattern is similar to the wild-type strain, the co-occurrence between the nervous system and general and others has increased from a Dice coefficient of 0.16 to 0.19. Furthermore, the Dice coefficients for the co-occurrence between the lower respiratory system and the nervous system, as well as between the lower respiratory system and the upper respiratory system, have increased from 0.15 to 0.19 and 0.18, respectively. In the Omicron variant, compared to the wild-type and Delta variant strains, the strongest co-occurrence is concentrated in the upper respiratory system. Specifically, the Dice coefficient between the upper respiratory system and the musculoskeletal system is 0.18, followed by the co-occurrence between the upper respiratory system and the nervous system, with a Dice coefficient of 0.15 (Figure 6).

**Fig 6.**
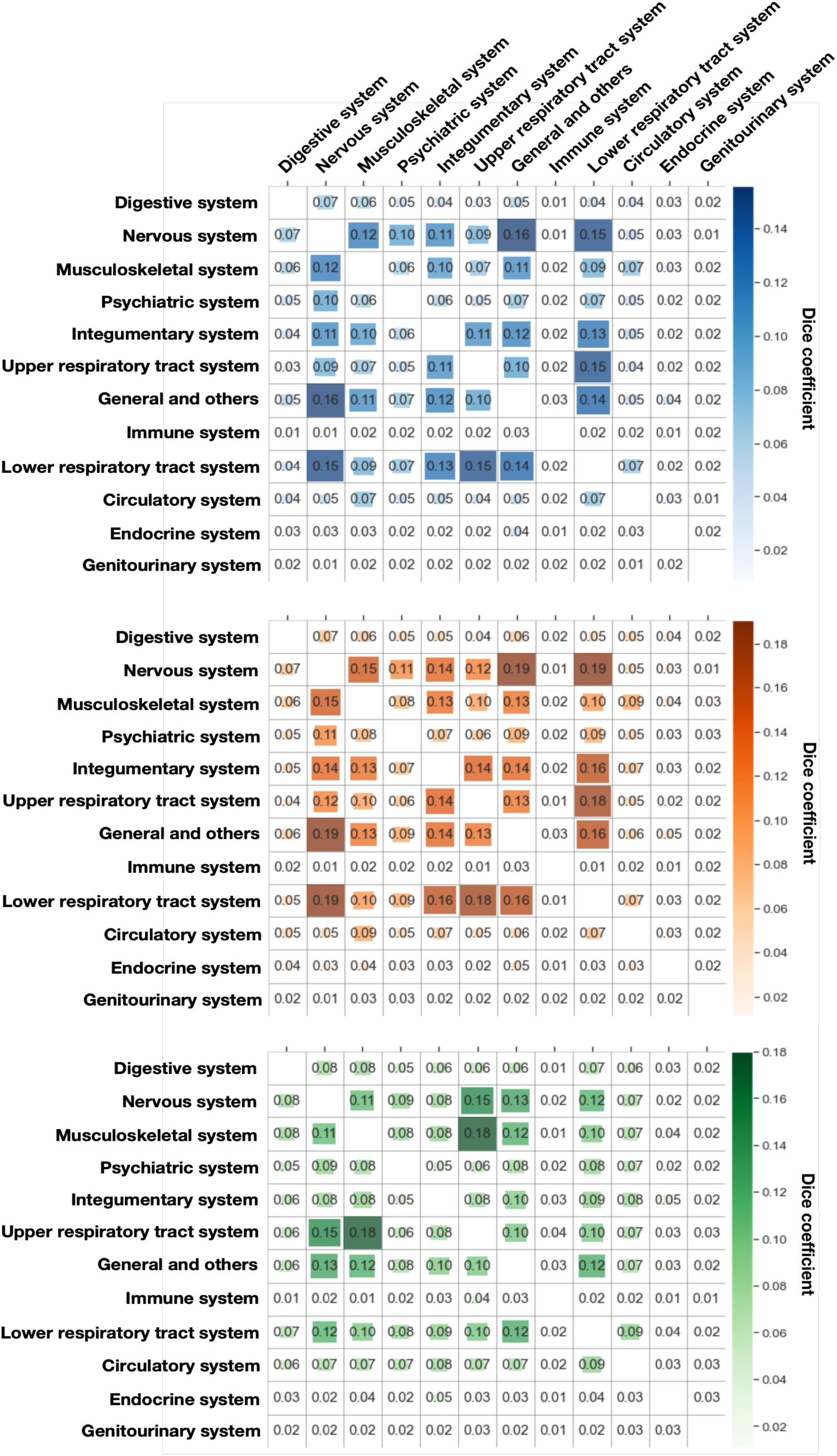
Symptom co-occurrence intensity among different variants. (Note: From top to bottom are the Wild-type, Delta variant, and Omicron variant, respectively.)

## Discussion

In this study, we developed a pipeline for denoising longitudinal social media data to monitor dynamic changes in disease symptom patterns, with COVID-19 as a use case. The pipeline begins with a rule-based NLP technique to identify self-reported symptom cases from large social media datasets and track their historical data over time. An NER model then extracts symptom information from this extensive text data. A USM classification model to identify the symptom mentions that reflects user’s health status. Our pipeline enables public health researchers to leverage vast amounts of noisy data from social media platforms to uncover the variability of disease symptom epidemiology.

There existing several studies[27,55,56] that have delved into identifying whether a user’s related symptom or disease is mentioned in a tweet, categorizing them into distinct classification tasks. For example, Luo et al [27] and Karisani et al [55] have annotated the symptom tweets with four labels: self-mention, other-mention, awareness and non-health, while Biddle et al [56] classified symptom tweets into three labels: figurative mentions, other mentions and health mentions. However, in the context of health monitoring tailored for a particular population, the primary objective is to extract information solely pertaining to the user’s own health status and does not require further segmentation of non-user-related-symptom. Consequently, only two labels were selected for our symptom tweets labeling: user-related-symptom and non-user-related-symptom. After denoising symptom tweets, we found that although a large number of tweets discussed symptoms, only about 39% of tweets containing symptom information reflected the individual’s personal experience. When analyzing symptom patterns in patients, it is critical to accurately identify the individual symptom experiences. This study shows that the frequency of mentions of symptoms such as headache, fatigue, and skin lesions increased after denoising with the USM model, while the frequency of mentions of chills and muscle cramps decreased. This suggests that the rate of user-related symptom of symptoms is related to the nature of the symptoms. One study [57] showed that about 44% of tweets containing health issue keywords disclosed user-related health status, but disclosure rates varied by health issue. For example, more than 80% of tweets about migraines and allergies were related to user themselves, whereas only 12% of tweets about abortion disclosed user-related information. Therefore, in the comprehensive prevalence analysis of different symptoms, denoising processing is extremely critical, otherwise the analysis results of some symptoms may be seriously biased and cannot truly reflect the actual prevalence trend.

The longitudinal analysis of symptom patterns showed that symptom prevalence across most physiological systems declined except upper respiratory tract systems during periods when different COVID-19 strains were predominant. During the pandemic dominated by different SARS-CoV-2 strains, the dynamic changes in symptom prevalence risk differ. This difference may be a result of different levels of immunity developed by natural infection or vaccination [58,59], the mutation of the viral spike protein leads to changes in the transmission ability, pathogenicity, and immune escape ability of the virus [60]. Specifically, we noticed that the Delta strain variant has a more persistent effect on the lower respiratory tract. This is in accordance with a previous study that shows that the Delta strain variant causes more severe and longer-lasting lower respiratory symptoms [61]. In addition, our finding that the Delta strain variant had a longer risk resolution period in the integumentary system is consistent with an online U.K. cohort study [62], which announced that cutaneous symptoms were more common and lasted longer in the Delta strain variant than in the Omicron strain variant.

We explored the symptom co-occurrence patterns caused by different SARS-CoV-2 strains and found that Delta variant and wild-type SARS-CoV-2 strains were highly similar in most symptoms co-occurrence, but the association between lower respiratory tract and nervous system symptoms was enhanced. Another study also used symptom co-occurrence network analysis to reveal similar symptom manifestations between gamma variant and wild-type strains [63]. In addition, previous studies have used co-occurrence network analysis of long-term COVID-19 patients to reveal complex relationships between symptoms, revealing that abnormal breathing, chest pain, and fatigue are related [53], which also suggests that respiratory dysfunction-related symptoms and neurological symptoms are more likely to co-occur. SARS-CoV-2 can invade host cells by binding to the Angiotensin-Converting Enzyme 2 (ACE2) receptor [64]. ACE2 receptors not only widely exist in the respiratory system, also distributed in the nervous system, they constitute the respiratory and nervous system symptoms concurrent biological basis. Furthermore, current scientific consensus believes that olfactory mucosa may be an important route for SARS-CoV-2 to enter the brain. The virus can use sensory nerve endings in this region to enter the brain through retrograde transport mechanisms, affecting multiple parts of the skull including the olfactory, trigeminal, and autonomic nervous systems [65]. This mechanism provides a possible explanation for the onset of neurologic symptoms, including loss of smell and taste, in patients with COVID-19. At the same time, the virus directly causes respiratory symptoms when it enters the respiratory tract and damages epithelial cells. Therefore, the dual involvement of the respiratory system and nervous system can also be regarded as a direct reflection of the unique biological characteristics of SARS-CoV-2 and its pathophysiological effects.

Our integrated pipeline shows promising results in monitoring the COVID-19 pandemic. This capability enables its application to the surveillance and analysis of other infectious diseases. By incorporating social media data, our pipeline enhances the monitoring of shifts in symptom patterns, thereby informing public health strategies. This approach not only keeps both the public and healthcare professionals informed about prevalent symptoms but also aids in anticipating the impacts of emergent viral variants of concern.

## Limitations

We acknowledge the limitations of our study. First, our analysis relied on self-reported positive cases of COVID-19 rather than confirmation by laboratory testing. Although we have adopted strict regular screening to ensure the accuracy of self-reported data as much as possible, it still cannot completely rule out the existence of false positive cases. Second, our strain groupings were based on SARS-CoV-2 strain prevalence data published by the CDC rather than on laboratory-tested strain genotyping, which may have included a small number of cases with other variants. Genotyping of laboratory-tested strains would help to distinguish the differences between different variants more precisely. Third, due to the privacy and security issues of social media data, it is difficult to cover variables such as age, gender, vaccination, and underlying diseases, and it is difficult to fully control confounding factors when analyzing the symptom patterns of different SARS-CoV-2 strains in this study. Nevertheless, the results of this study are generally consistent with the conclusions of several clinical case studies. Finally, as with other social media-based public health studies, this study suffers from potential sample bias problems because the demographic characteristics of the social media user group do not fully reflect the distribution of the overall population. This limitation may have affected our assessment of the representativity of reporting COVID-19 cases and their symptoms.

## Conclusion

We developed an integrated pipeline for denoising longitudinal social media data to monitor the evolution of symptom patterns during the pandemic over time. By applying this pipeline to up to two years of COVID-19 related social media data, we enabled retrospective tracking and analysis of the health status of a substantial cohort of self-reported COVID-19-positive patients. Our analysis revealed notable variations in symptom patterns across different SARS-CoV-2 strains. This pipeline not only provides valuable insights into COVID-19 symptomatology but also establishes a robust framework for epidemic monitoring, crucial for addressing current and future public health challenges.

## Data Availability

All data produced in the present study are available upon reasonable request to the authors.

